# Impact of lock down relaxation on the COVID-19 epidemic trajectory in Bangladesh

**DOI:** 10.1101/2020.07.20.20158527

**Authors:** Shafiun Nahin Shimul, Mofakhar Hussain, Abu Jamil Faisel, Syed Abdul Hamid

## Abstract

In this projection exercise, we analyzed the circumstances of the COVID-19 pandemic in Bangladesh and used multiple methods to characterize the epidemic curve. We merged several publicly available data sets for the purpose. Projections using actual Government data as of June 16, 2020 reveals that the epidemic curve for Bangladesh may be different from that of developed countries and quite similar to such curves in countries in the region. This is true, both in terms of incidence of cases (total number of cases per million population) and length of the epidemic (months to peak or flatten the epidemic curve). We find that while Bangladesh went into lockdown early, efforts to maintain lockdown at a national level was relaxed and new cases accelerated; with significant growth happening since lifting of lockdown on May 31. Our estimates indicate prevalence of COVID-19 may be between 200,000 and 600,000 towards end of the year, may take 9 months (270 days) to flatten the epidemic curve, lifting of the lockdown may have increased total cases by 60 to 100% and may have prolonged the epidemic by additional 2-3 months.

## Introduction

Coronavirus (COVID-19) is a new strain from the coronavirus family and was first detected in the Wuhan province, China, in December, 2019. By mid-February, 20,000 infected cases were in found China and by beginning of March, about 80,000 were infected with about 4,000 deaths. By early March, about 20,000 people in Europe were infected with about 1,000 deaths. On January 30 the World Health Organization (WHO) declared COVID-19 as a Public Health Emergency and then on March 11, WHO declared COVID-19 as a global pandemic. (1)

Researchers have determined that COVID-19 is a highly infectious virus that may spread person to person in multiple ways(2) and the rate of transmission often represented by the reproductive number or R_0_ was estimated to be higher than known infectious virus (2). It was also found that 1-6% of those infected may die (3). As of the writing of this paper there is no cure or vaccination for COVID-19.

Since the early days of the COVID-19 pandemic, researchers have warned that unless mitigated, the dramatic increase in infected cases may overwhelm health systems. Thus, it became critical that estimation exercise of the potential number of cases be undertaken for Bangladesh, to inform timing and levels of potential cases. This paper reviews the current status of COVID-19 pandemic in Bangladesh, discusses different policies undertaken to mitigate the spread, evaluates and estimates total spread of COVID-19 cases in Bangladesh using multiple methods.

## Background

Disease dynamics of COVID-19 indicate that transmission may happen from droplets to nose and eyes and may go from infected person to surface or to other persons (2). Once a person is infected, there is an incubation period where infected person does not show any symptoms. This incubation period may be between 5 and 14 days. However, the infected person, who does not have symptoms, may be able to transmit the virus (4). Hence, while symptomatic persons may infect others, it is also possible infected asymptomatic persons may also spread the disease (5). It is for these reasons that face masks, hand washing hygiene and social distancing has become so important in containing the spread of the disease.

Most infected with COVID-19 do not show any symptoms and those that do have fever, cough, fatigue, slight dyspnoea, sore throat, headache and conjunctivitis (4). The course of the infection is mild for most infected; thus 80-90% recover without any serious impacts; however, about 10% may have trouble breathing and low blood oxygen level and require hospitalization. A further 5% may experience life threatening condition such as respiratory failure and multiorgan failure. Thus, case fatality rate, defined as percentage of infected patients who may die, for COVID-19 range from 2-5% and this percentage increases with age and for those with chronic comorbid conditions such as diabetes, asthma and heart disease. Thus spread of COVID-19 in Bangladesh poses a significant threat; and without active intervention by Government, even if a small percentage of the population, say 5%, are infected, this translates to 8,000,000 needing hospitalization and over 200,000 deaths.

Observing the spread and potential significant health effects, many researchers estimated potential impact of COVID-19 pandemic to help policy makers prepare respective countries and their systems to face this pandemic. The main thrust of these estimations has been to encourage policy makers to implement strong Non-Pharmaceutical Intervention (NPI) such as social distancing, hand hygiene and wearing of face masks, self quarantining, all to reduce the potential spread of the pandemic (6). Research has also found that testing and isolating or quarantining infected persons may mitigate the spread of the infection (7).

These estimations use epidemiologic models such as the Suspected-Infected-Recovery (SIR) model to project the number of infected cases, number of deaths and potential demand for health care, specially hospital ICU use during times of the peak of pandemic (8). Subsequent studies focused on the impact of NPI on the spread of the disease. One such study was by Ferguson et al. published on March 16, 2020 (9). Without strong NPI, the study estimated that R_0_, which is a measure of rate of infection transmission, could be about 2.4 and about 80% of the population would be infected in UK and US and about 2.2M people would die in USA alone. With NPI and other mitigation efforts, the study noted that health care system would be overwhelmed and deaths in US would reach about 1.1M to 1.2M people. Less than two weeks later, a study by Murray et al (10) predicted that even with strong NPI estimated deaths over a 4-month period would be about 81,000 in the US. A Canadian study estimated initial infection rate of 56% in Ontario, Canada and NPI could reduce that infection rate to 2%(11). A recent European study estimated initial reproduction number for COVID-19 is 3.8 and NPI has had significant impact on the spread of the disease resulting in reproduction number of less than 1 at 0.86 and final infection rate of about 3.2-4% of the population (12).

In the context of Bangladesh, the first COVID-19 case was identified on March 9 and by March 23, 2020 total cases reached 33. It is at that point that the government of Bangladesh announced shutdown^1^ of public, private offices, schools and colleges from March 26,2020 for 10 days (15). Subsequently all public transport systems were shut down (16). Police, the Army and other security forces were deployed to enforce social distancing. Announcement of the initial lockdown, in the form of a general holiday, for 10 days was followed by multiple extensions. Since the announcement on March 24, 2020, there have been several occasions where there have been relaxation of lockdown including allowing garments workers to join work(17), allowing stores to open and relaxation of travel during Eid religious holiday(18). Finally, the Government announced that all general holiday will end on May 30 and work and travel can resume on May 31; however, there would be a strict enforcement of NPI such as social distancing, and vulnerable population such as the elderly, pregnant women and those with multiple chronic diseases are discouraged to re-join their workplace. Schools and colleges were to remain closed.

In the mean time the Government has attempted to increase capacity of the health care system, including increased number of COVID-19 testing sites from 2 to more than 50 (19), testing about 15,000 a day; recruitment of 2,000 new doctors and 6,000 nurses (20) and opening 2000+ beds field hospitals (21).

In the second week of May, cyclone Amphan, with a land speed of 185km/h, made land fall in the coastal region of Bangladesh. The cyclone left a trail of destruction including 28 deaths, destructions of 200,000 homes and damage to about 200 bridges and culverts (21).

To mitigate economic impact of the pandemic and subsequent cyclone, the Government has implemented multiple stimulus packages targeted at workers, small and large business, garments and agricultural sectors, low income/informal workers and those affected by cyclone Amphan. The Asian Development Fund (ADB), the International Monetary Fund (IMF) and the World Bank has pledged aid and loan (23) to support Bangladesh in this crisis.

Studies on Bangladesh specific COVID-19 models estimated that by end of May, 2020 about 89 million may be infected and total deaths could be 500,000.(13). Another study estimated that without NPI, daily new cases at peak of the epidemic would be nine Million and deaths over 18 months would be over 200,000 and with NPI, new daily peak would be 36,000 and cumulative deaths would be about 5,000 (14).

Although studies have attempted to understand the impact of various interventions and project cases using real data and established models, there has not been any study of COVID-19 projections for Bangladesh that estimates impact of policy changes using multiple models.

In light of these events, in this paper we attempt to estimate the epidemic curve for Bangladesh under alternative scenarios using multiple methods. Pre-post analysis is done to evaluate impact of the lifting of the lockdown on May 31. The significant contribution of this paper is to apply models from diverse fields: epidemiology, econometrics, and data analytics (machine learning tools) which allowed us to capitalize the strength of various fields. Since there is an insurmountable amount of unknown that follow COVID-19, using various tools can provide a stronger prediction. This paper uses actuals figures as of June 16,2020 obtained from IEDCR, official Government authority to provide COVID-19 data. At the time, about 100 days had passed since the first case and total number of infected cases in Bangladesh was about 90,000.

## Methodology

As noted before researchers employ standard epi models to predict levels of infections in a pandemic. However due to lack of Bangladesh specific parameter data on contact and transmission rates and significant measurement issues with available data, standard method of SIR may not be sufficient for projecting cases in Bangladesh. Second, unknown characteristics of COVID-19 creates substantial uncertainty and finally changing nature of the NPI in Bangladesh since the beginning of the epidemic requires multiple approaches to projecting cases.

### The SIR method

One method is the Susceptible-Infected-Recovered (SIR) model developed by Kermack, and McKendrick (24). The model uses parameters of incidence, transmissibility, duration of infections and recovery/deaths to estimate trajectory of incidence of infection and deaths over time. Note recovery here means those who have recovered or died as result of the infection and are removed from those who are susceptible.

The SIR model is used to estimate rates of infections for highly infectious disease. Often termed as compartmental model, the model relates changes in suspected case (S), infected cases (I) and recovered or death (R) via 3 differential equations as shown below:

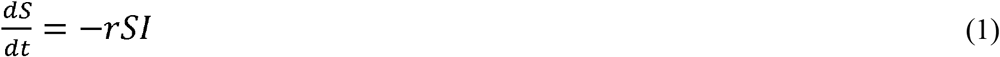

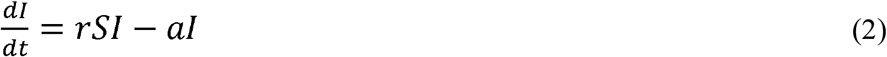

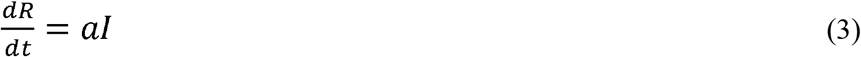

where r represent transmission rate and a represent rate of recovery or death among infected. Note R+S+ I= population, which is assumed to be constant and for Bangladesh it is assumed to be 161 Million.

Equation (1) describes change in suspected cases is inversely related to rate of contact (S*I) and rate of transmission; equation (2) describes change in infected cases depends on number of newly infected (*rSI*) minus number of cases infected that are removed *al*; equation (3) describes change in recovery or death depends on rate of recovery (a) and number of infected cases I.

Equation (2), can be rearranged to represent R_0_ = r S/a, where r/a represent contact ratio or fraction of the population that comes in contact with persons who are infected during the period of the epidemic and R_0_ reproduction number. R_0_ value of greater than 1 represents a epidemic as number of newly infected will be greater than number of currently infected.

Challenges with the SIR model is that all parameters are subject to local conditions; so, contact rate may be affected by labour market decisions of individuals, recovery rate may be affected by availability of local hospital capacity etc. These conditions vary widely country to country.

Variation of the SIR model was developed by Battista (25) and it is that model that is used in this study.

### The Gompertz method

A method to simulate the expected exponential growth of COVID-19 cases in a S-shaped curve can also be estimated using a Gompertz. The simplest form of the Gompertz curve can be expressed as:

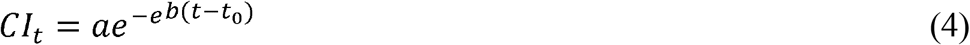

CI_t_ is the cumulative cases at time t, a is the projected number of maximum cases, b is the growth rate of infection, *t_0_* is the start date. The cumulative cases are driven by the three parameters; a, b and *t_0_* and using regression techniques these may be estimated.

Gompertz curve has been previously used to estimate patterns of human height growth as well as spread of an epidemic (26,27,28). The main advantage of using Gompertz distribution is that it allows having a longer tail which is a likely case for COVID-19. In addition, this distribution is flexible in parameters compared to say logistic and exponential models.

### The Quadratic Method

This approach is to fit actual data to a quadratic (parabolic) function that reflects the epidemic curve observed in countries that have experienced the COVID-19 pandemic over a given number of days. The first incidence in Bangladesh was in March 8, full 3 months after the first incidence in China. That means daily data was available for all countries for 90+ days to analyze experience in other countries with the virus.

The approach is to use estimation of new case count based on quadratic function where the dependent variable is a function of square of the dependant variable. Using this approach, number of new infections in a country *i* at time *t* after *x* days of the onset of the infection can be estimated using the following formulae.

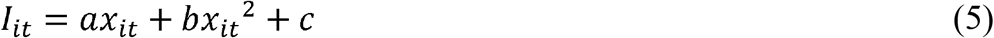

In this model, number of daily cases in a country is assumed to depend on the number of days since COVID-19 cases reached 100 (x) and its squared value. It is assumed that *x* capture epidemies measures such as contact rates and transmissions rates and its square values assumes the epidemic will peak in a country after certain days in the country. It is also important to note maximum level of *I* when the first differential of equation 4 is set to zero and maximum value of x is reached when 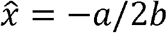. A useful aspect of this method is that it allows us to change the values of a or b to decrease or increase the days over which the epidemic will reach a peak. Thus higher values of a or lower values of b will increase the days to peak of the epidemic. These adjustments to parameters may be informed by evidence of impact of NPI or lack there of.

Using regression methods, parameters a, b and can be estimated and these parameters can then be applied to futures *x* for specific country to calculate *I* for the future.

### The Facebook Prophet method

Another method is an open sourced time series forecasting tool developed by Facebook known as Prophet (29). This platform has previously been used in COVID-19 (30). The algorithm automatically selects changes in data points, runs piecewise linear or logistic regressions accounting for variations in data due to various events, say weekdays. Another advantage of this tool is that it can capture recent changes and can provide better forecast taking both old and relatively newer growths. This tool has been extremely popular among the data scientists and machine learning experts, and some famous machine learning sites provide projections using this tools, such as in Katana ML firm (https://katanaml.io/). The prophet model can be run using R or python. We applied python fbprophet library to produce our results.

### Pre-Post Analysis

Although recommended approach to manage a pandemic is to implement NPI strategies until pandemic has reached a peak, many developing countries do not have the economic capacity to sustain strong NPI measures for a lengthy period across the country. As a result, many countries are forced to relax their respective NPI enforcement even when daily cases have not peaked. Bangladesh is one such country where NPI strategy was lifted on the 99th day since the first case, even though new case counts were on the rise.

Since each of these models are based on actuals as of a specific date, estimation based on two different times can be used to evaluate impact of NPI policy pre and post lifting of NPI.

Thus impact lifting of NPI is D*_i_* for country i can be calculated as

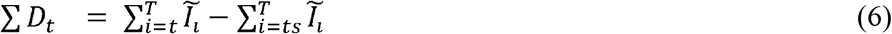

Assume NPI was relaxed on date t and current date is ts, where ts is greater than t, T is the when total cases reaches a peak (new cases approach zero) then impact of NPI is estimated to be the difference in projected total case using actuals as of date t versus actuals as of ts. In such as case, 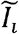 is the actual or estimated incidence of total cases at time *i, ts* is the date on which NPI was implemented and *t* is the date of the most recent actuals and *T* is the final date of the of the projection period. Note that *t* is greater than *ts*.

### Data

Country level annual demographic, health care related data and income was obtained from World Bank’s ‘Health and Nutrition Program’ data (31) and data included percentage of population over 65, percentage of GDP spent of health care, life expectancy at birth; per-capita income at purchasing power parity (PPP) was used as per-capita income data. All data points from the World Banks was for 2016. To compare Bangladesh with other countries categorization was necessary and one categorization developed was per-capita income. With this categorization Bangladesh can be compared with other countries of similar per-capita income. For this approach, countries are ranked according to per-capita income, and all countries were grouped into 5 buckets (quintiles); such that about 20% of the bottom per-capita income countries are put in group 1, 20% of subsequent ranked per-capita income countries are put in group 2 and so on.

Another categorization was done to compare Bangladesh to others in the region. It was assumed other countries in the region may have similar demographics, per-capita income, health care capacity as a result managing COVID-19 may pose the same challenges and result in similar outcome. Thus India, Pakistan, Vietnam, Thailand, Philippines, Nepal, Bhutan, Indonesia, Malaysia, Sri Lanka were included in this category.

Country level COVID-19 related daily data was obtained from three different sources. Europeans Union’s European Centre for Disease Prevention and Control (ECDPC) data provides country level daily cases and deaths for COVID-19 (32). The country specific daily test data was downloaded from the “github” site(33). Country specific daily reduction in cell phone traffic was downloaded from Google mobility (34). Google publishes data on location specific mobility data by type of location for each country and by date. The report shows percentage change in cell phone traffic in a given location in a country or region for a given date compared to a baseline of Jan-3 to Feb-4 of 2020. The location identified include retail, grocery, parks, transit, workplace and residential. Review of the mobility data for Bangladesh showed that data for parks and grocery were varying during the lock down period, workplace data was deemed to not reliable as significant portion of the working population is in the informal sector (thus no specific workplace location); finally trends in transit and retail data were the same. Thus for the purpose of this study we use changes in cell phone traffic in transit location in Bangladesh. It is assumed this trend reflects changes in social distancing and other NPI, where lower levels (higher negative levels) reflect stronger social distancing and vice versa.

Country level daily data from these 3 sources and annual figures was then merged to create a single data set for this study.

### Results

For comparative purpose, we pursue a cohort type analysis where we compare measures for Bangladesh to similar measures of other selected countries where all countries were in the 99^th^ day of their respective COVID-19 epidemic.

There about 100 countries selected for comparison. The category created to represent countries per-capita income is INC_RNK_CAT. The data shows Bangladesh is in INC_RNK_CAT =1 among all the countries ranked by Per Capita Income. All measures are reported as median in their respective group. Bangladesh has lower spending on health than its comparators (2.5% vs 5.5%), however in terms of life expectancy, Bangladesh does better (71.8 vs 62.7). Age distribution data shows Bangladesh has more elderly as *%* of all population compared to its cohort; however this percentage is lower in Bangladesh than other countries with higher per capita income.

These aggregate measures appear to show as per capita income increases, countries tend to spend more of their income on health, they appear to have higher life expectancy and as a result percentage of the elderly population increases.

COVID-19 related measures show that transit mobility has decreased by 37% as of June 16 in Bangladesh compared to baseline mobility of January/February,2020. This is within range of the same measure of other countries; however other countries may have different epidemic experience and thus the decline will need to be evaluated in the context of experience of each country. Tests per million population show Bangladesh has higher rates compared to it cohort(93 vs 85) and lower than rates in higher income countries. COVID-19 total cases per million for Bangladesh appear to be significantly higher than its cohort countries (562 vs 285); note this is measured 100 days after the date of the first case for all countries. Thus Bangladesh appear to have much higher rate of infection compared to other countries in same income groups. For deaths per million, rates for Bangladesh is higher than its cohorts and in terms of case fatality (which equals total deaths divided by total infected), rates for Bangladesh is comparable to its cohort.

Compared to high income countries, Bangladesh appear to have lower cases per million; this may be a reflection of lower levels of testing or other factors. WHO has provided guidance on testing which is based on test positivity rate. The positivity rate is the ratio of the number of people who test positive for COVID-19 to all those who are tested. As testing programs scale up, the number of tests go up and if the virus is contained or on its way to be contained, the number of people testing positive go down, thus the positivity rate is expected to go down. WHO has advised that testing programs should try to test such that positivity rate is between 3 and 12%. Currently, In Bangladesh, the positivity rate has been about 20%. Thus in Bangladesh testing may need to be at-least doubles to reduce positivity rate. There could be other reasons that results in lower cases in Bangladesh which may include levels of infection spread and different levels of immunity to the virus.

Higher income countries appear to have higher levels of testing, levels of cases and deaths and higher levels of case fatality. High case fatality in high income countries could be explained by the fact that these countries have more elderly population and more of the elderly population are institutionalized. A single outbreak in one of these institutions, which typically houses 100-200 seniors, may result in significant outbreak and a large number may of the seniors may not survive. Thus recent evidence show that most of the elderly deaths (those 65 years or older) were not in the community but in long-term care facilities. About 80% of the deaths in Canada and 40% of the deaths in USA were in long-term care facilities (39).

In table 1B, we compare Bangladesh with 10 other regional countries. Several observations are relevant: health spending in Bangladesh is the lowest, life expectancy and elderly as percentage of the population are within range of its neighbors. In terms of COVID-19 measures, Google’s Transit data show Bangladesh had higher values than only two of neighbors Vietnam and Pakistan and for two different reasons. By the 99^th^ day Thailand was registering less than 50 cases a day and Nepal was registering less than 5 cases a day. Vietnam has already reached its peak daily cases and thus was opening up its economy while Pakistan was still experiencing exponential growth in daily cases as is Bangladesh. Total cases data indicate Bangladesh has the highest rate of infections among its neighbors. The rate of deaths in Bangladesh is second highest, highest being that of Pakistan.

**Table 1A:**
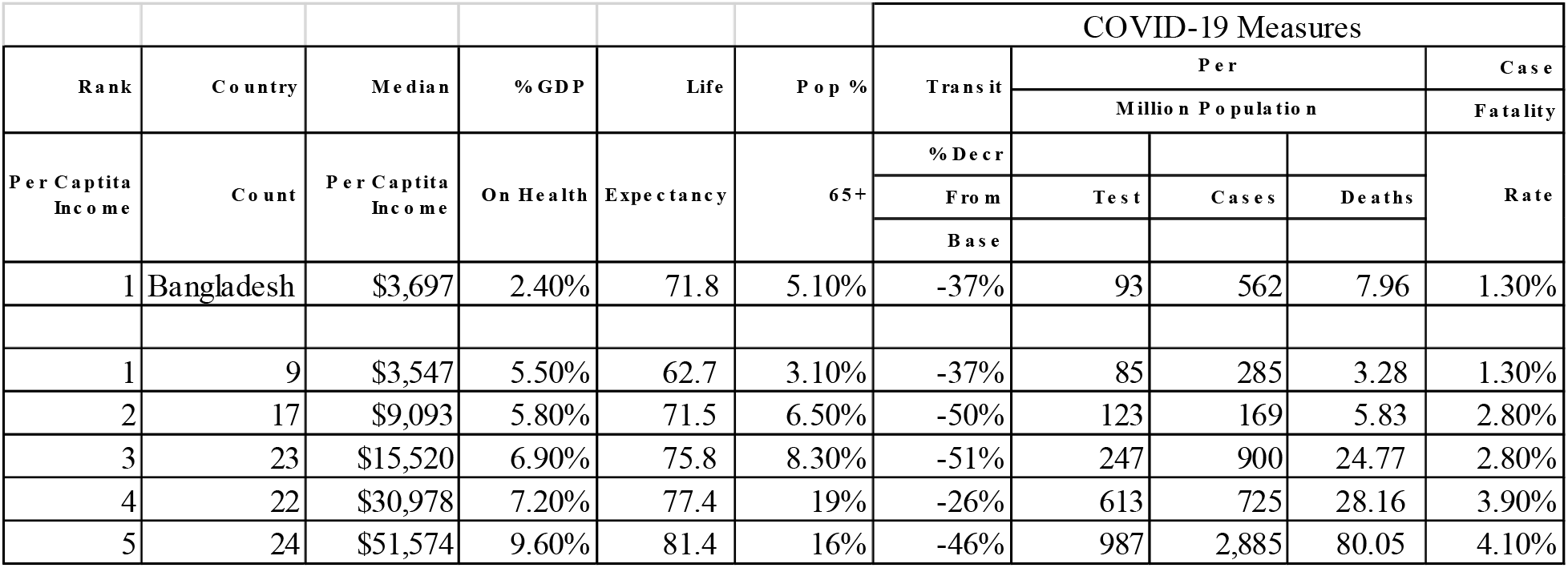
Comparison of Bangladesh with other countries ranked by Per Capita Income As of the 99^th^ day from first case date.

**Table 1B:**
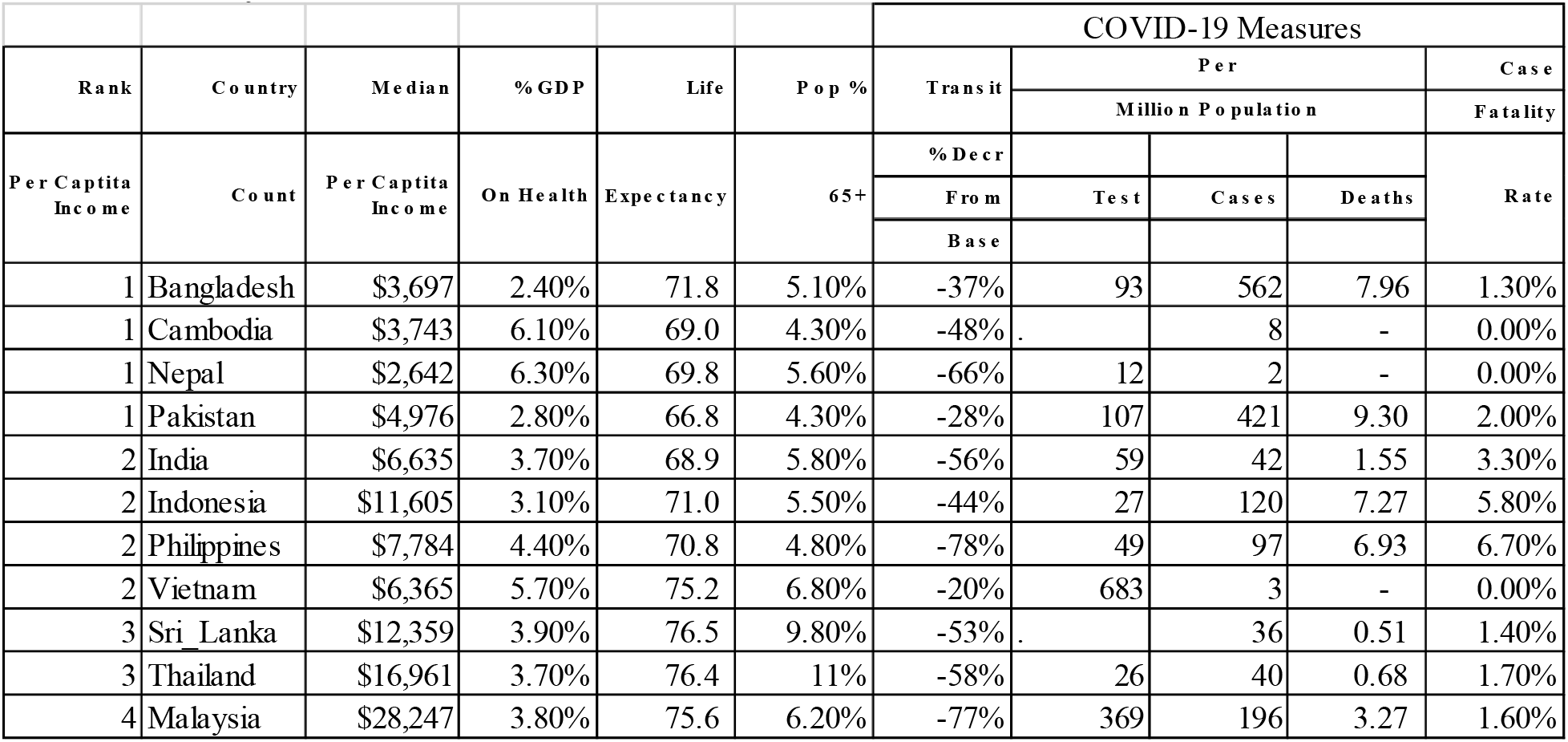
Comparison of Bangladesh with other countries in the region. As of the 99th day from first case date.

Table 2 shows Bangladesh specific weekly data for transit, tests, new and total cases. Noting that Bangladesh announced a general holiday on march 28, 2020 with an attempt to increase levels of NPI, it appears NPI was most effective in Bangladesh in around middle of April when the percentage decline in transit mobility was highest at around 70% but then mobility has been increasing and the most dramatic increase coming in on first week of June with almost 20% increase in mobility. Data also shows significant increases in testing and new cases since mid-April from about 2,000 tests to about 15,000 tests a day and new cases increase from 300 to about 3,000 cases a day. Thus tests per day has increased 7 folds while new cases have increased by 10 folds.

**Table 2:** COVID-19 Weekly Measures

| Week Of Year | Last Day of Week | Daily Transit % Change From Baseline | Daily Tests-Max | Daily New Cases-Max | Daily Total Cases-Max |
| --- | --- | --- | --- | --- | --- |
| 1 | March 14, 2020 | 0% | 0 | 0 | 0 |
| 2 | March 21, 2020 | -19% | 49 | 7 | 17 |
| 3 | March 28, 2020 | -70% | 106 | 9 | 48 |
| 4 | April 4, 2020 | -68% | 434 | 5 | 61 |
| 5 | April 11, 2020 | -73% | 1,184 | 112 | 424 |
| 6 | April 18, 2020 | -73% | 2,190 | 341 | 1,838 |
| 7 | April 25, 2020 | -71% | 3,686 | 503 | 4,689 |
| 8 | May 2, 2020 | -71% | 5,827 | 641 | 8,238 |
| 9 | May 9, 2020 | -65% | 6,244 | 790 | 13,134 |
| 0 | May 16, 2020 | -56% | 8,582 | 1,202 | 20,065 |
| 1 | May 23, 2020 | -62% | 10,834 | 1,773 | 30,205 |
| 2 | May 30, 2020 | -62% | 11,291 | 2,523 | 42,844 |
| 3 | June 6, 2020 | -46% | 14,088 | 2,911 | 60,391 |
| 4 | June 13, 2020 | -45% | 16,638 | 3,471 | 81,523 |
| 5 | June 16, 2020 | -37% | 15,038 | 3,141 | 90,619 |

Source: ECDPC, Github and Google

**Table 2B:** Parameter and Key Estimates: SIR Model

|  |  |  | Actual as of |  |
| --- | --- | --- | --- | --- |
|  |  |  | Jun-16 | May-30 |
| Number of observations |  |  | 94 | 77 |
| Initial Infected |  |  | 96,699 | 45,942 |
| Contact frequency |  |  | 0.49 | 0.524 |
| Removal frequency |  |  | 0.423 | 0.442 |
| Infectious period |  |  | 2.4 | 2.3 |
| Final Suseptibles |  |  | 550,053 | 222,864 |
| Final Infected |  |  | 194,527 | 92,574 |
| Total epidemic duration |  |  | 292 | 233 |
| Basic Reproduction number |  |  | 1.159 | 1.183 |
| Reproduction number |  |  | 1.013 | 1.018 |
| End reproduction number |  |  | 0.856 | 0.836 |

**Table 2C.**
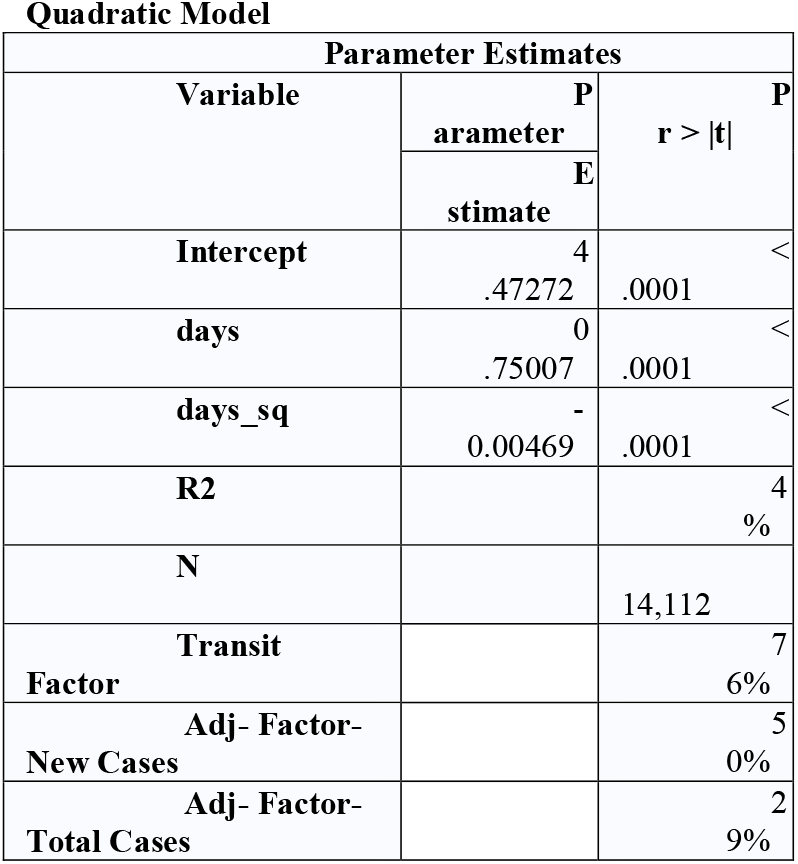
Parameter Estimates: Quadratic Model

In Bangladesh, national lockdown was announced less than 20 days after the first case was identified. Many developed nations had sub-national lock downs and national lock downs was announced much later than that in Bangladesh. In Canada and United States, for instance, sub-national lock downs were announced and varied by Province/State. In these countries there was never a national lock down. In UK and Italy, national lockdown was announced long after the date of the first case. In Italy for instance first case was detected in January 31 and the national lockdown was announced on March 9, 38 days later (35),. In the region Pakistan, India and Malaysia announced national lock downs well after the first case was detected. In Malaysia targeted national lock down was announced about 50 days after the first case (36) and India national lock down was also announced 50 days after the first case (39). As the transit mobility data Graph 1 and Graph 2 shows, Bangladesh experienced a significant drop in mobility well before other countries and the reduction was significant to about -70%. That reduction remained in place for about 40 days after which mobility tended to increase signaling decreases in NPI compliance. In most cases, this trend contrasts with other countries where the reduction remained in place for first 100 days.

**Graph 1:**
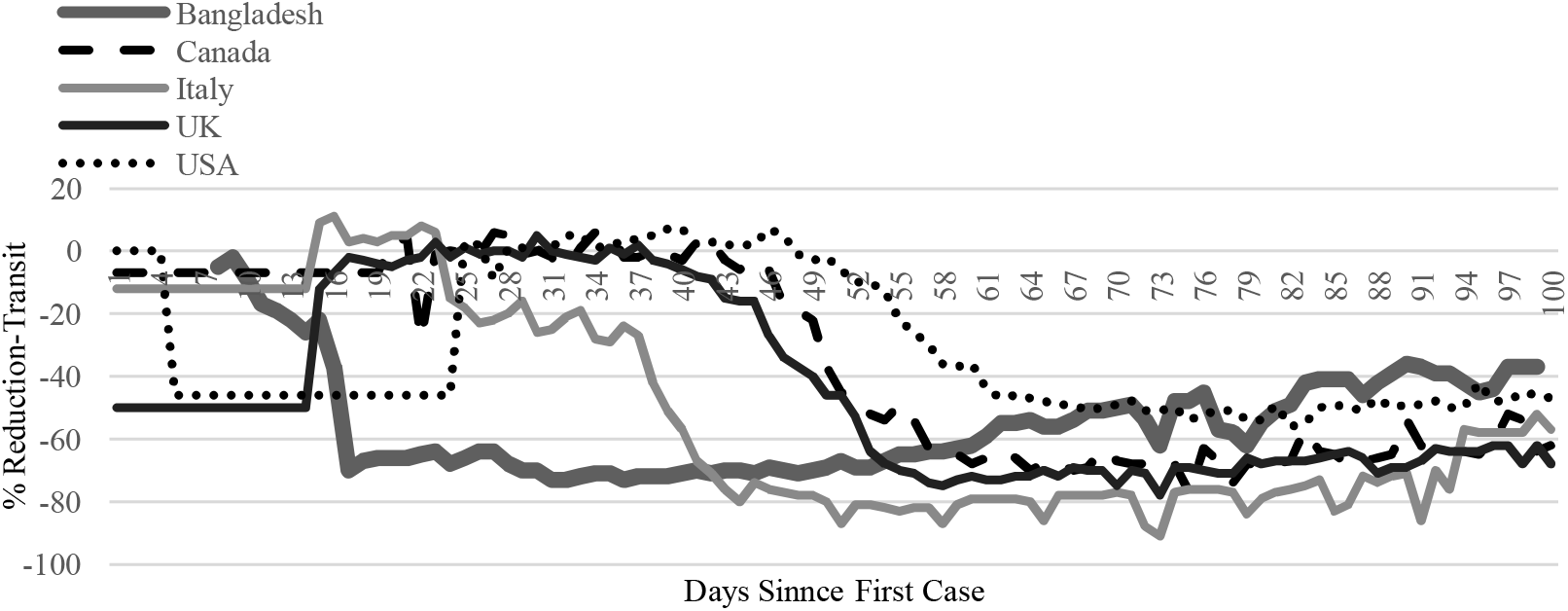
Change in Transit Data by Days since First Case: Bangladesh and Other Developed Countries

**Graph 2:**
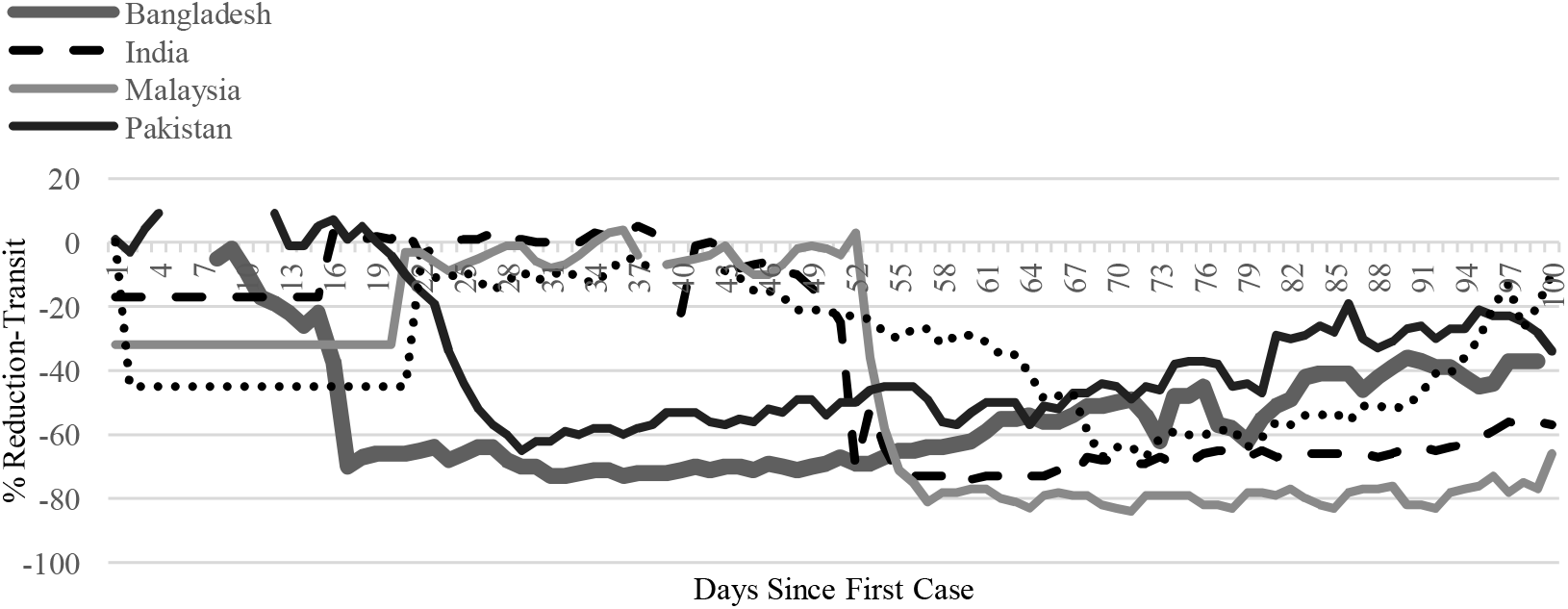
Change in Transit Data by Days since First Case: Bangladesh and Other Regional Countries

Trends in new cases per day per million population (Graph 3 and Graph 4) indicate that the cases in Bangladesh is still growing. When compared to regional countries, the levels in Bangladesh is significantly higher than other countries in the region. More importantly, this trend appear to be accelerating, where acceleration may have started between the 70^th^-80^th^ days. This translated to calendar days between middle of May to end of May. Thus the announcement of lifting of lockdown on June 1 coincides with acceleration of new cases. It is for this reason that we use May 30 as the cut-off point for pre-lockdown analysis. It is expected that projected number of cases in the future will be significantly higher if we use actuals cases as of June 16 (post lockdown) versus projection using actuals thru May 30 (pre-lockdown). Further in certain models, the change in mobility between May 30 to June 16 can be used as proxy for reduction of NPI.

**Graph 3:**
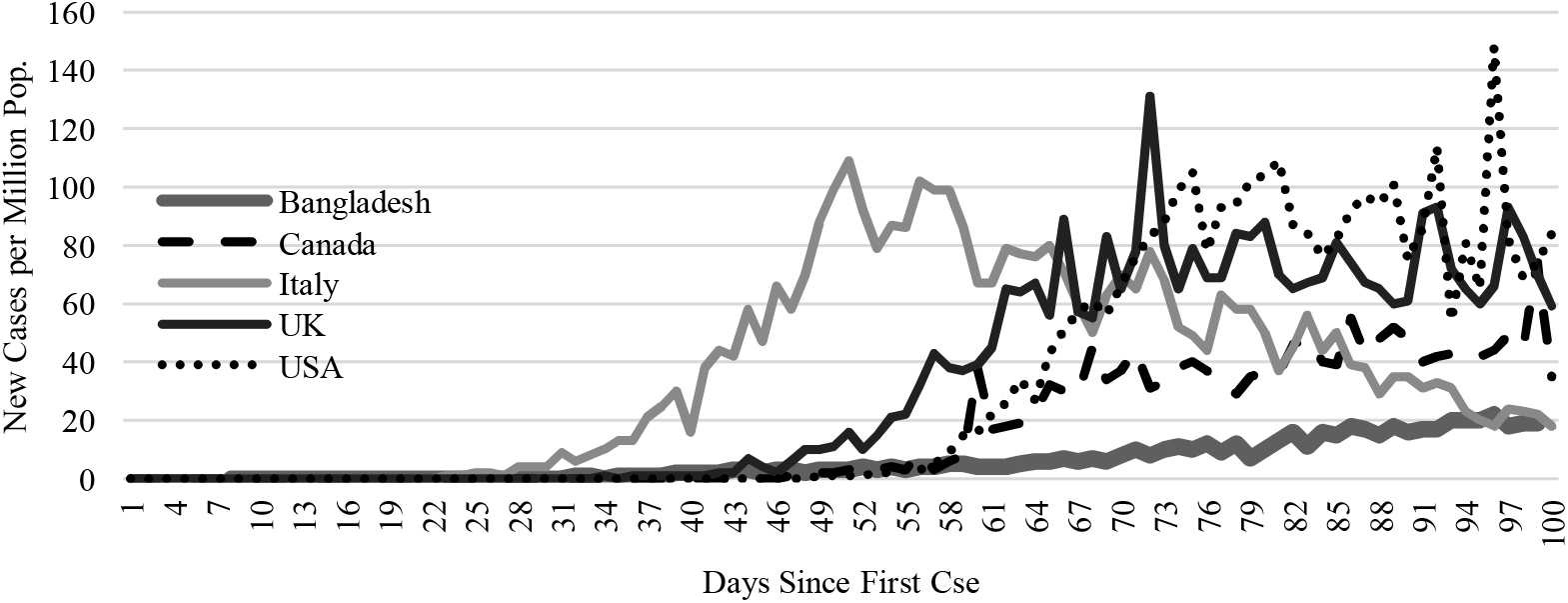
New Cases Per Day Per Million Population Bangladesh and Other Developed Countries

**Graph 4:**
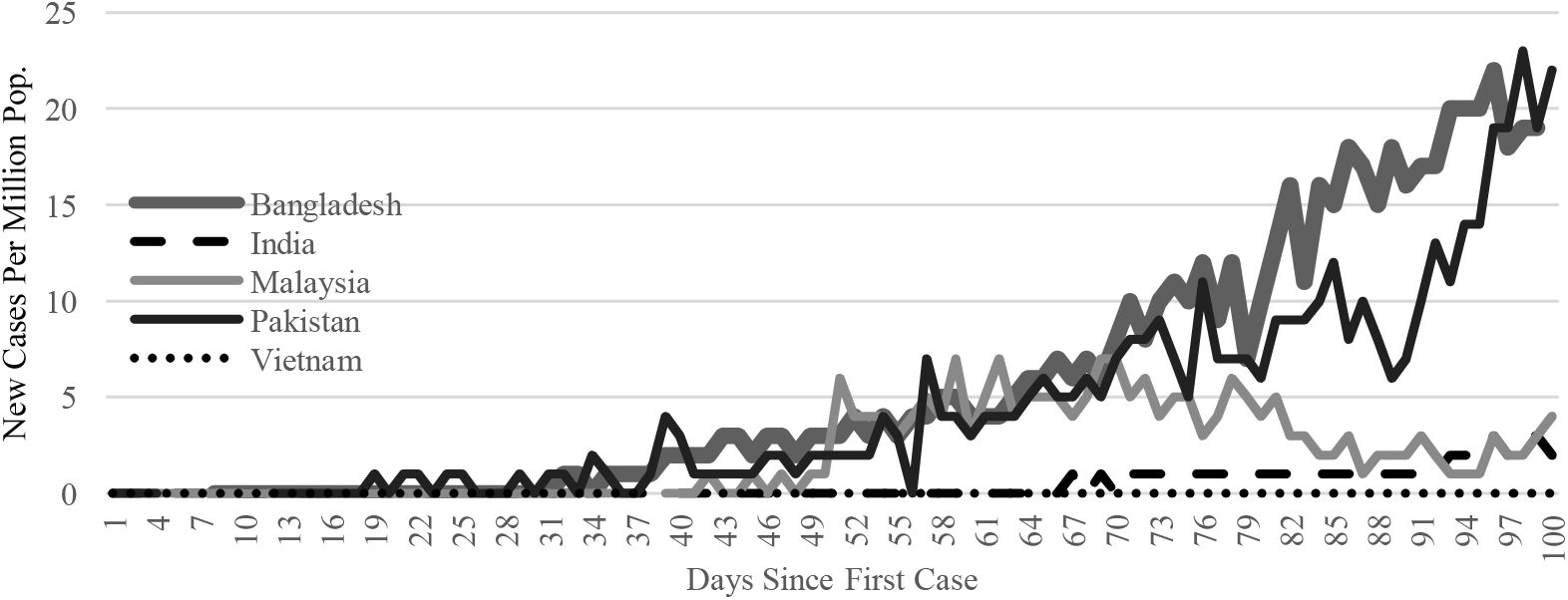
New Cases Per Day Per Million Population Bangladesh and Other Developed Countries

### Projection

Actual new and total cases as of June 16, 2020 were used for this study. As of that date, the epidemic has been in country for about 99 days since the first case and as of June 16, 2020 there were about 90,000 cases with most recent moving 3-day average daily cases of about 3,100. The ‘pre’ period was defined as the period from date of first case to May 30, 2020 or about 80 days. As of May 30, 2020, there about 43,000 total cases and daily new cases was about 2,500.

#### SIR Model

Estimation using the SIR methodology, using actual cases as of June 16,2020, indicates that new cases will peak in around the middle of June and total cases will peak at about 194,527 and early phase of peaks in total cases will be reached around August, 2020. The epidemic is projected to end in December, 2020. The model assumes a contact rate of 0.49, recovery/removal rate of 0.423, infection period of 2.4 days. The model reports a reproduction number or R_0_ of 1.013 and epidemic period of 292 days.

For the ‘pre’ period, the SIR model estimates total cases to peak at 93,000 in October, 2020. The model assumes a contact rate of 0.524, recovery/removal rate of 0.442, infection period of 2.3 days. The model reports R_0_ of 1.018 and epidemic period of 233 days.

Thus most recent actual cases projects that the epidemic will continue to the end of the year, reach about 194,000 cases. The lock down relaxation that came into effect first of week of June may have resulted in doubling the number of total cases and extend the epidemic by about 60 days.

Using the Gompertz method, projection results in 609,333 total cases by September, 2020. Daily new cases are estimated to peak around early August at about 6,500 a day.

#### Quadratic Model

Under the ‘quadratic’ modelling approach, regression is run using equation 4 above. In this approach country level daily new cases per million is regressed on country specific time measures such as days since 100^th^ case and its square. The resulting parameters are then adjusted to account for increased transit activity in Bangladesh using Google mobility data. The relationship between Google mobility data and COVID-19 cases was explored by Yilmazkuday (2020) who showed a strong relationship between mobility data and new cases. In the context of Bangladesh the transit data showed as May 30,2020 the percentage change from baseline was about -62% and that percentage increased -37% by June 16. Thus mobility increased by about 24%. This evidence is used to lower the value of a in equation 4 by multiplying by 0.76 and recalculating the number of projected new cases. These projected new cases are then further adjusted to align with actual levels using factors noted in Table x.

Regression results indicate that coefficient of days and days_sq have the expected sign. This means that number of new cases increases with days post the first 100 days and negative sign on the days_sq coefficient implies that new cases reaches a maximum on a specific level value of day. Under the base regression, the estimated number of days to reach maximum of new daily cases is about 90 days (.75/(2*0.004)=93.75). Bangladesh is yet to achieve daily maximum; plus recent relaxation of lockdowns is likely to extend the daily maximum longer. So to adjust for that prospect, the coefficient was adjusted by the transit factor of 0.76.

As Graphs 9 and 10 shows, the quadratic method shows that new cases is projected to peak around July-2020 at about 3,600 a day and total cases likely to peak around November-2020 at about 310,000 total cases. If the current case fatality holds to the end, total deaths is estimated to be about 4,000. Given that there is wide-spread community spread, even with the recently announced efforts of Red Zoning, total cases may continue to increase. Projected fatality levels may be under-estimated as current fatality numbers may be under reported and future case fatality rate may increase as a result of health care system will come increased pressure and will struggle to save lives.

**Graph 5:**
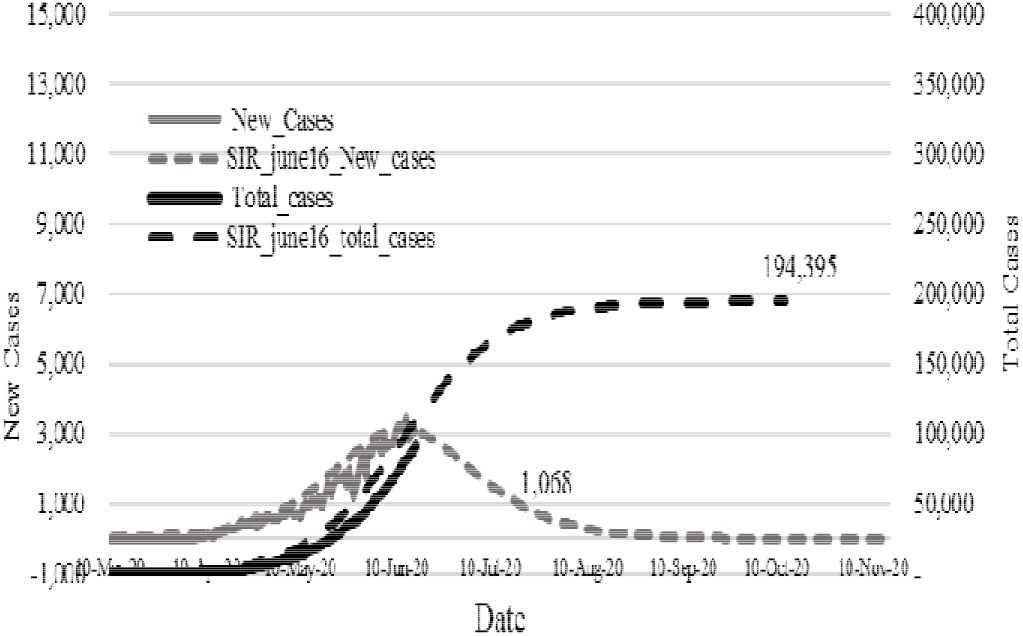
Epidemic Curve, Bangladesh Actuals as of June 16, 2020 Method: SIR

**Graph 6:**
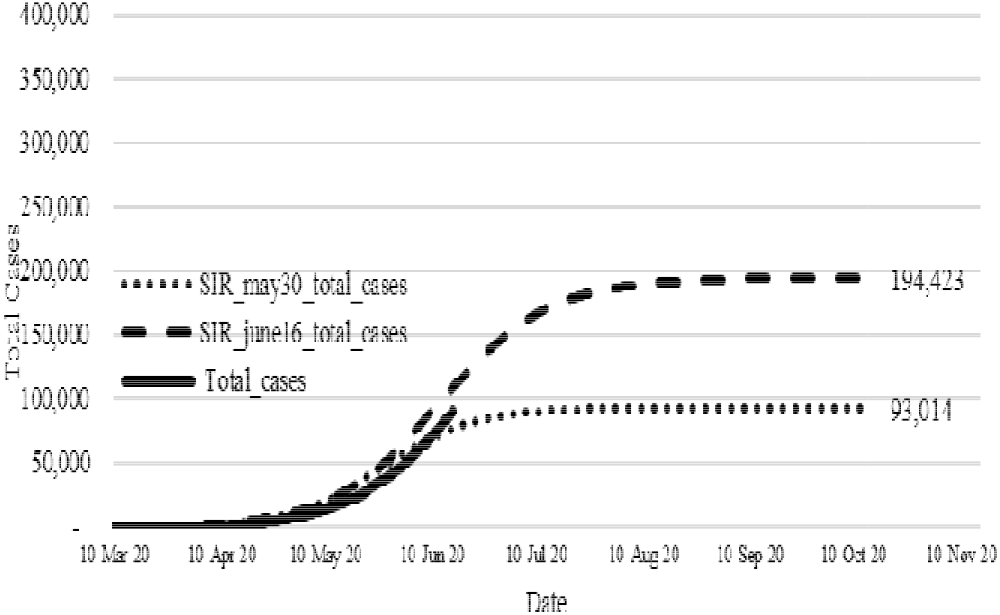
Epidemic Curve, Bangladesh Pre(Actuals as of May 30) vs Post(Actual as of June 16, 2020) Method: SIR

**Graph 7:**
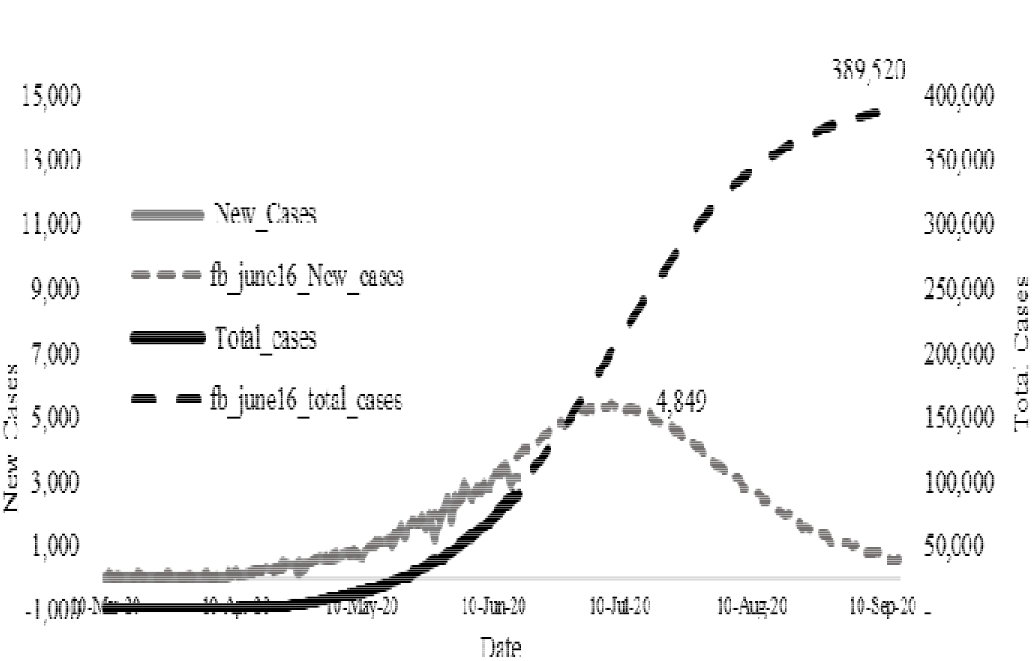
Epidemic Curve, Bangladesh Actuals as of June 16, 2020 Method: Compertz

**Graph 8:**
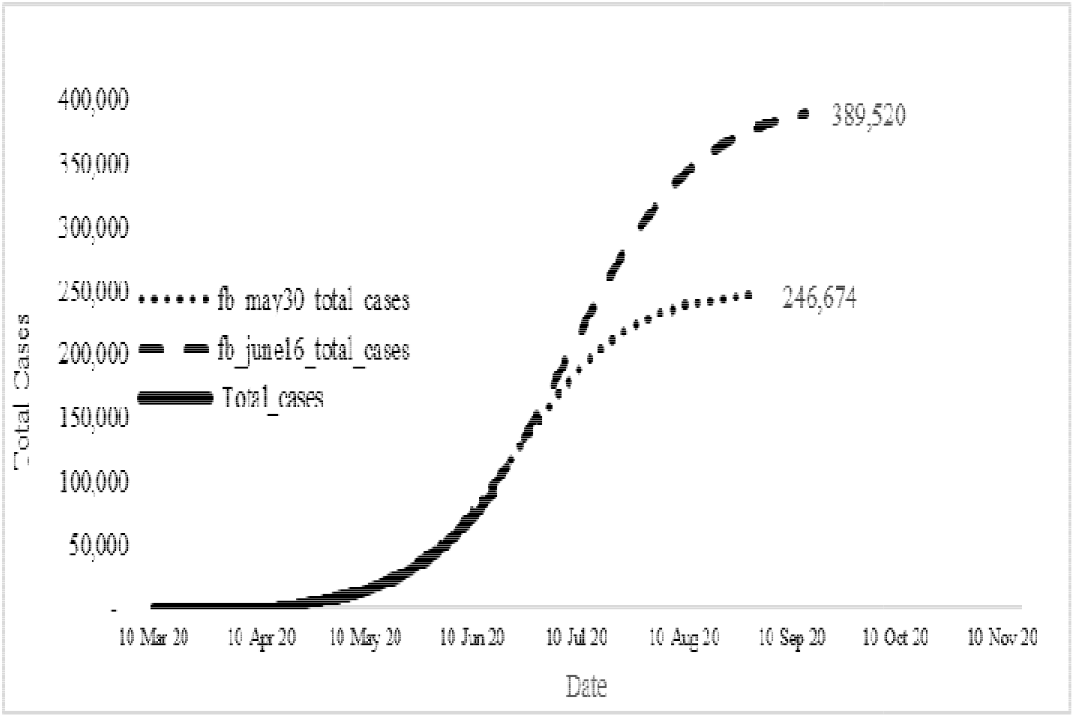
Epidemic Curve, Bangladesh Pre(Actuals as of May 30) vs Post(Actual as of June 16,2020) Method: SIR

**Graph 9:**
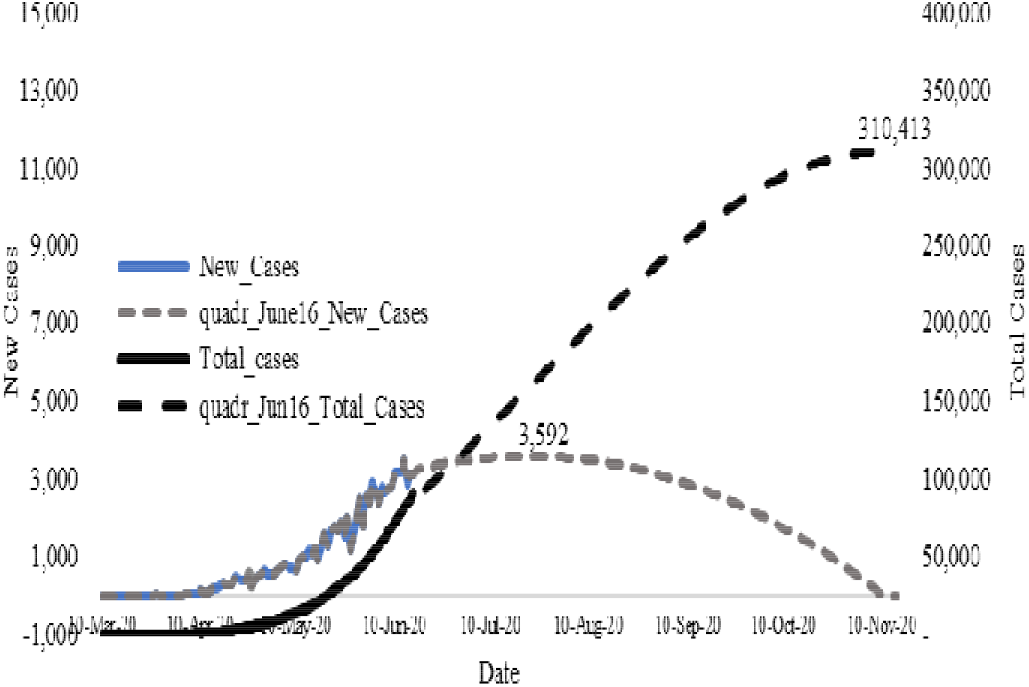
Epidemic Curve, Bangladesh Actuals as of June 16, 2020 Method: Quadratic

**Graph 10:**
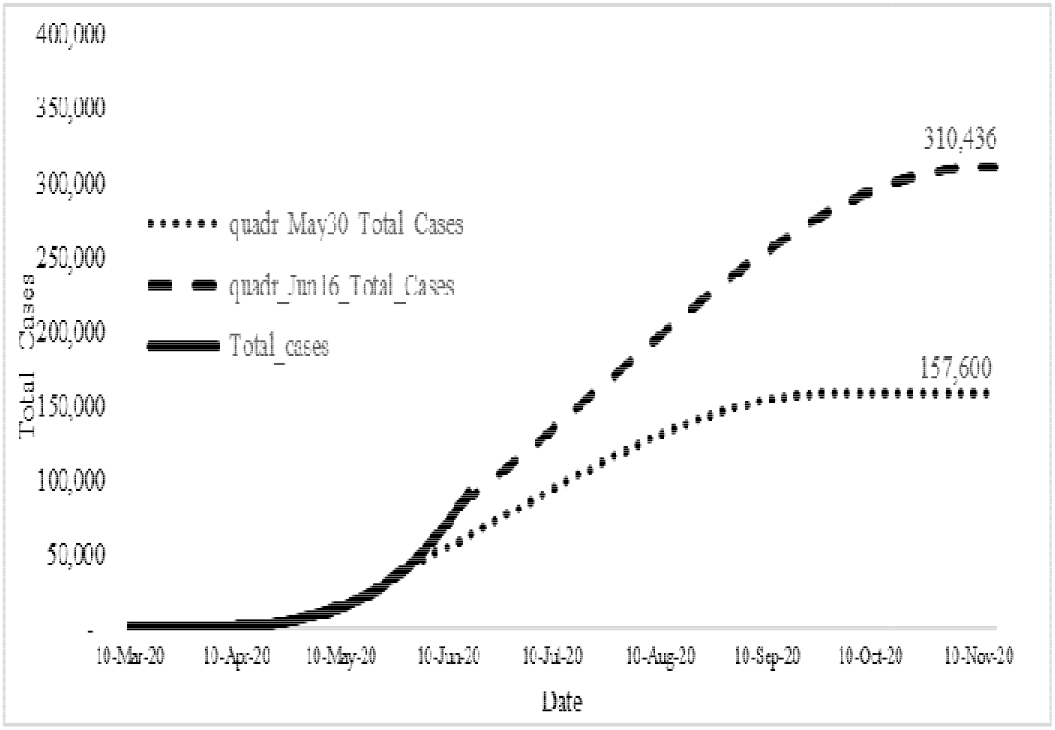
Epidemic Curve, Bangladesh Pre(Actuals as of May 30) vs Post(Actual as of June 16,2020) Method: Quadratic

#### Facebook Prophet Model (FB)

The Facebook Prophet approach results in daily cases to peak at about 4,800 around first week of July and total cases to peak around 388,000 in first week of September. Projected daily and total cases tracks very close to actual levels.

Using actuals as of May 30, the projection results in total cases to peak around the same time in September but the number of total cases would be significantly lower at about 247,000.

Comparison across all 4 models, as shown in Table 3, reveals that total cases may peak between September to November and levels of cases may range from 194,000 to 615,000. Using the most recent case fatality rate and assuming same rates remain throughout the epidemic, total deaths may range from 2,600 to 8,200.

**Table 3:**
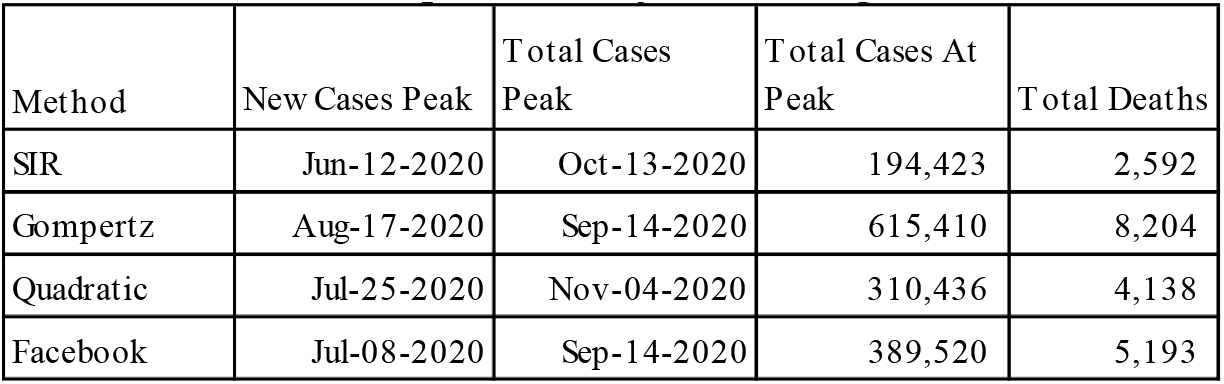
COVID-19 Epidemic Projection-Bangladesh

Comparison of NPI impact across the 4 methods is reflected through the difference between projected cases as of June versus end of May, shown in Table 4. With NPI, projected cases would be between 93,000 to 322,000; thus lifting of NPI may have resulted in increase of COVID-19 cases by 58% to 109%.

**Table 4:** COVID-19 Epidemic Projection-Bangladesh

| Method | Pre-Estimation | Current Estimation | NPI Impact | NPI Impact |
| --- | --- | --- | --- | --- |
|  | Total Cases |  | # | % |
| SIR | 93,014 | 194,423 | 101,409 | 109% |
| Gompertz | 322,412 | 615,410 | 292,998 | 91% |
| Quadratic | 157,600 | 310,436 | 152,836 | 97% |
| Facebook | 246,674 | 389,520 | 142,846 | 58% |

### Policy Engagement

The outcome of these analytic exercises was shared with key public officials for considerations and they have been using it consistently. So far the researchers have provided to the Health Government officials in the form of Technical Brief. The last one being # 10 and this process is on going. Appendix A provides example of such a report.

It was proposed that such analysis would assist key policy and decision makers in charge of managing the COVID-19 pandemic. After several iterations, it was decided that the results of the SIR model and district level data highlighting areas of improvements and concerns would be created.

## Discussion

All models projects that the current COVID-19 model may peak around the time frame towards end of the year. Total number of cases have a significant range multiple of 3, from about 200,000 to about 600,000; although 3 of the models predicts total cases to be 390,000 or lower. The high range may reflect the uneven growth of cases since the start of the epidemic. Rate of infection spreading ranges from R_0_ of 1.006 to 1.02.

The trajectory of COVID-19 epidemic in Bangladesh also reflects a much longer period of infection, here the daily peak is expected to be in July, 4 months after the first case was detected. NPI impacts supports the idea that lifting of the NPI has had significant impact in increasing potential cases from about 60% to 100% higher.

This study has several limitations. The use of multiple models to predict the outcome of an epidemic in a single country may result in outcome that varies significantly from one another, thus increasing the level of uncertainty. Limited data availability of Bangladesh specific transmission and recovery rates forces us to select initial values that may not be appropriate. Time specific influential events such holidays, major climate events and subnational events could not be captured in any of the models. Some of these events may extend the epidemic period. This study also does not take into account recent changes in policy such as assignment of red zone, lock down of neighborhoods, increases in contact tracing and other measures that government has taken to mitigate the spread of the virus. This study does not address the direct and indirect health effects of the COVID-19 epidemic. Future analysis could address these shortcomings.

There may also be a need to better understand and apply measures not directly captured in epidemiological reporting; we may need to include sociological and anthropological considerations in localities to better address the appropriateness of measures and methods in our modelling approach (37).

## Conclusion

Using multiple models, this study estimates the trajectory of the COVID-19 pandemic in Bangladesh. Results show a wide variation in the total number of cases may range from 200,000 to 600,000 towards the end of the year. This paper also shows that NPI polices have significant impact on the intensity and length of the epidemic; suggesting that lifting of NOI may have extend the epidemic by 3 months and increase prevalence between 60 to 100%. Substantial uncertainty exist as the rate of transmission of the virus is unknown, public compliance with NPI efforts has been mixed and governments ability to enforce and sustain NPI policies is challenged due to the negative effects they have on economic life.

## Data Availability

Data will be available upon request.

## Appendix A: Technical Briefing #8

June 16, 2020

Estimation of the number of potential infected cases as a result of the COVID-19 pandemic in Bangladesh^2^

Note: IEDCR data (national level) used as of June 16, 2020^3^; since district level data is not reliable as they stand, no district level analysis is done

### A. Purpose

Purpose of this document is to provide technical background behind the COVID-19 case projections for Bangladesh.

### B. Projection

This work tries to use publicly available data of IEDCR to estimate the total number of cases over the coming days for Bangladesh

### C. Methodology & Results

Several modeling approaches are used to understand the pattern and to make projections. These are:

✓ SIR model
✓ Modeling using forecasting tools developed by Facebook analytics team.

### D. Findings

#### D.1. Findings from SIR MODEL

SIR Model: We used the real data provided by IEDCR to estimate the parameters required for SIR model, and the following graphs provides the results from that model.

##### Interpretation of the findings

**Top graph:** Indicates the actual and projected cases. The top graph is illustrated with three colours. Pink color indicates the danger zone and infection will continue until we reach yellow zone when growth expected to steady for while until it reaches in the green zone i.e. the graph is reaching plateau. This graph says that if the current rate continues, our infection will grow until the end of July, then it may get stable (grow but at lower rate) for about three to four weeks until early September and then slowed down. Epidemic will continue until the end of the year, at least.

**Bottom graph:** The bottom shows the growth factors. We would like to the region where the growth factor is less than 1, which may not happen until the end of July, if the current rate continues.

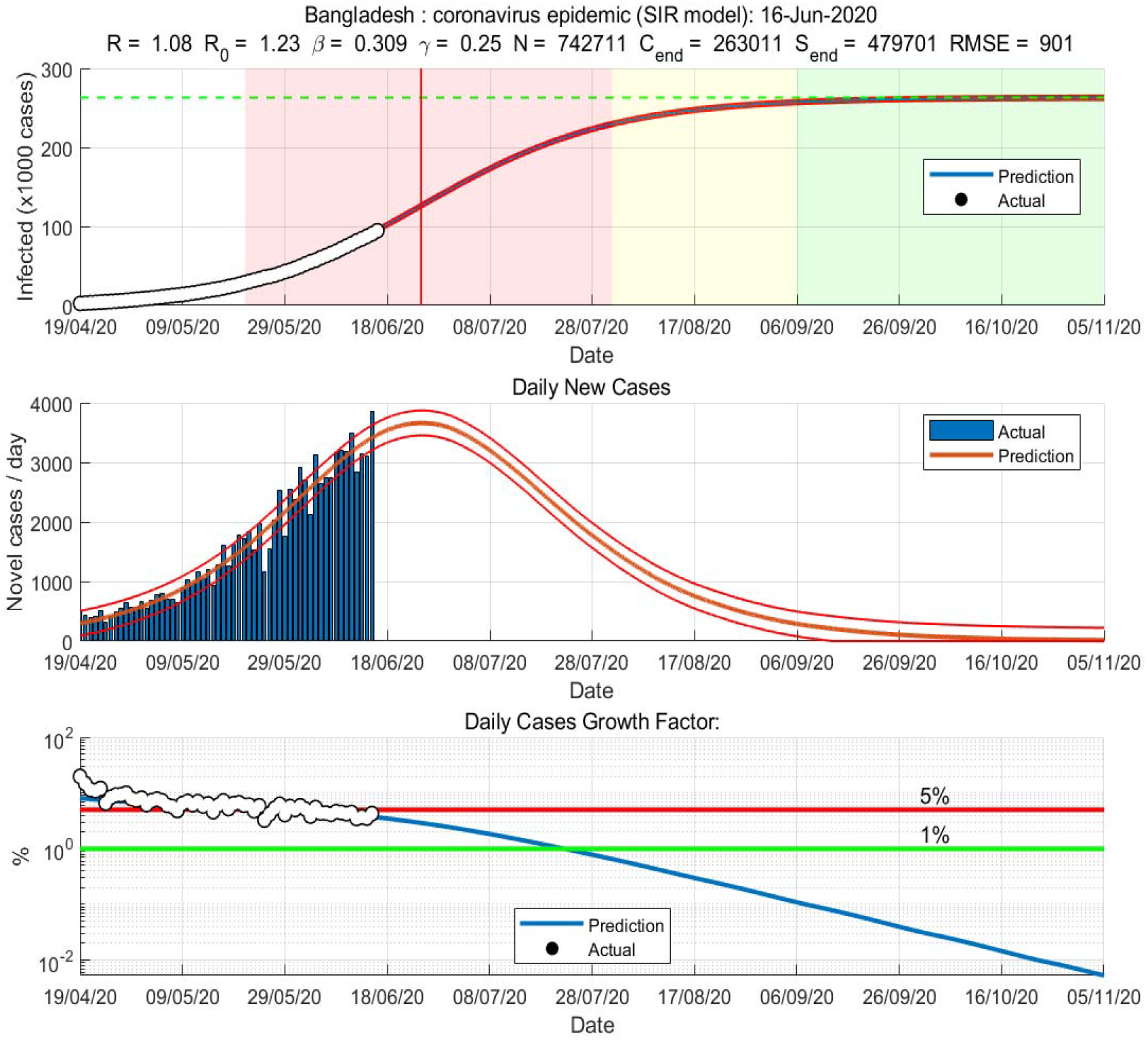

**Bangladesh:** coronavims epidemic (SIR model): 16-Jun-2020

#### D2. Findings from Time Series and Forecasting Tools of FB

The detail findings of time series model and SIGMOID Curve (Logistic and Gompertz Distribution) are not shown here. Gompertz distribution is less restrictive in terms of value of parameters it can adopt and so this is very likely situation. SIR model seems to draw same conclusion as the results from Gompertz distribution-so it is not reported. Facebook Analytic Tool is mostly like case given that everything is open now, and actual cases might be even higher. This model predicts the we may reach at pick by mid July with 5000-6000 daily new cases, however, it may even shift to the right with more number of cases. The following line charts provides likely scenario of Bangladesh. Right hand size axis shows total cases, and the left side axis shows new cases. Purple line shows predicted new cases, and organize bar shows predicted total.

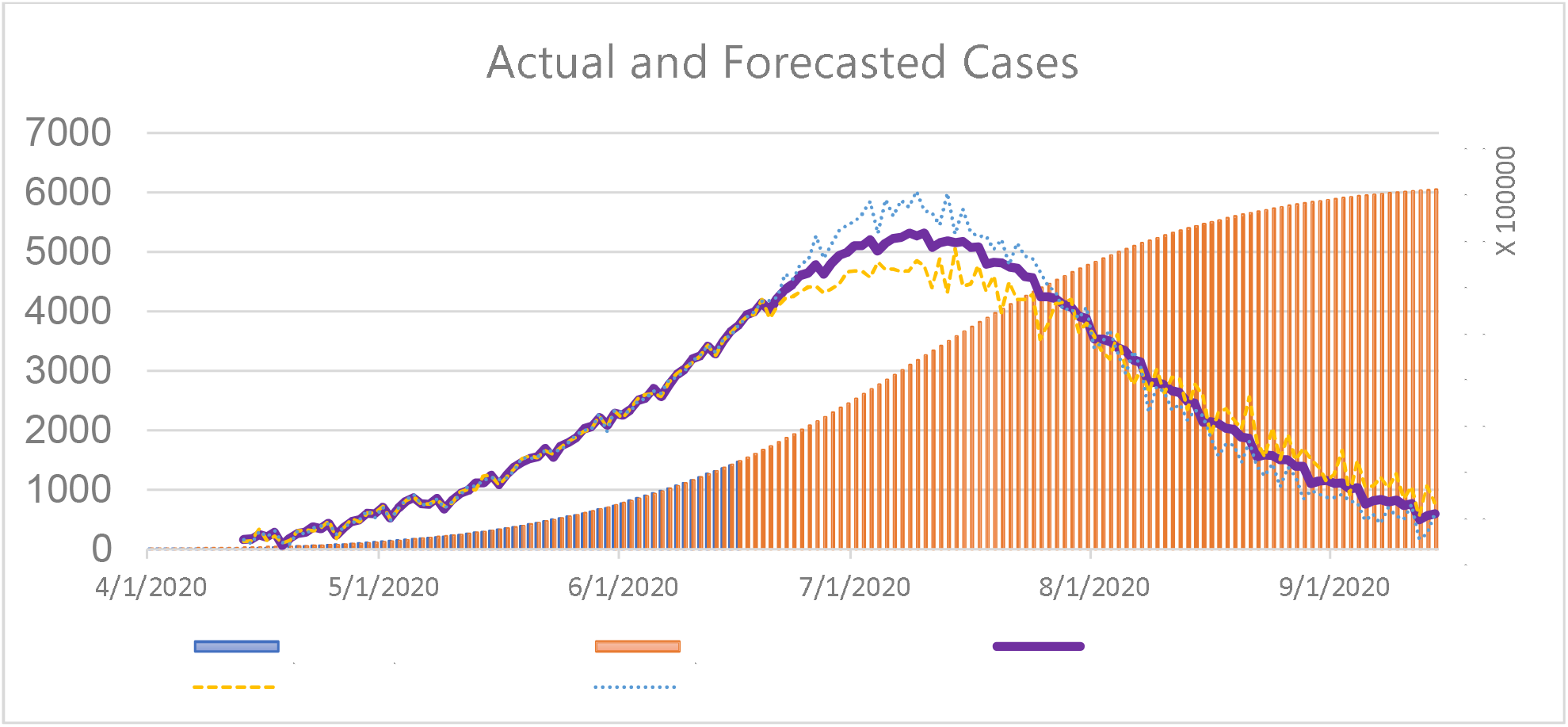

### E. Impact of economy opening

Since the economy has opened, we have more contacts and so the rate of transmission has also increased. This can be easily seen from the analysis

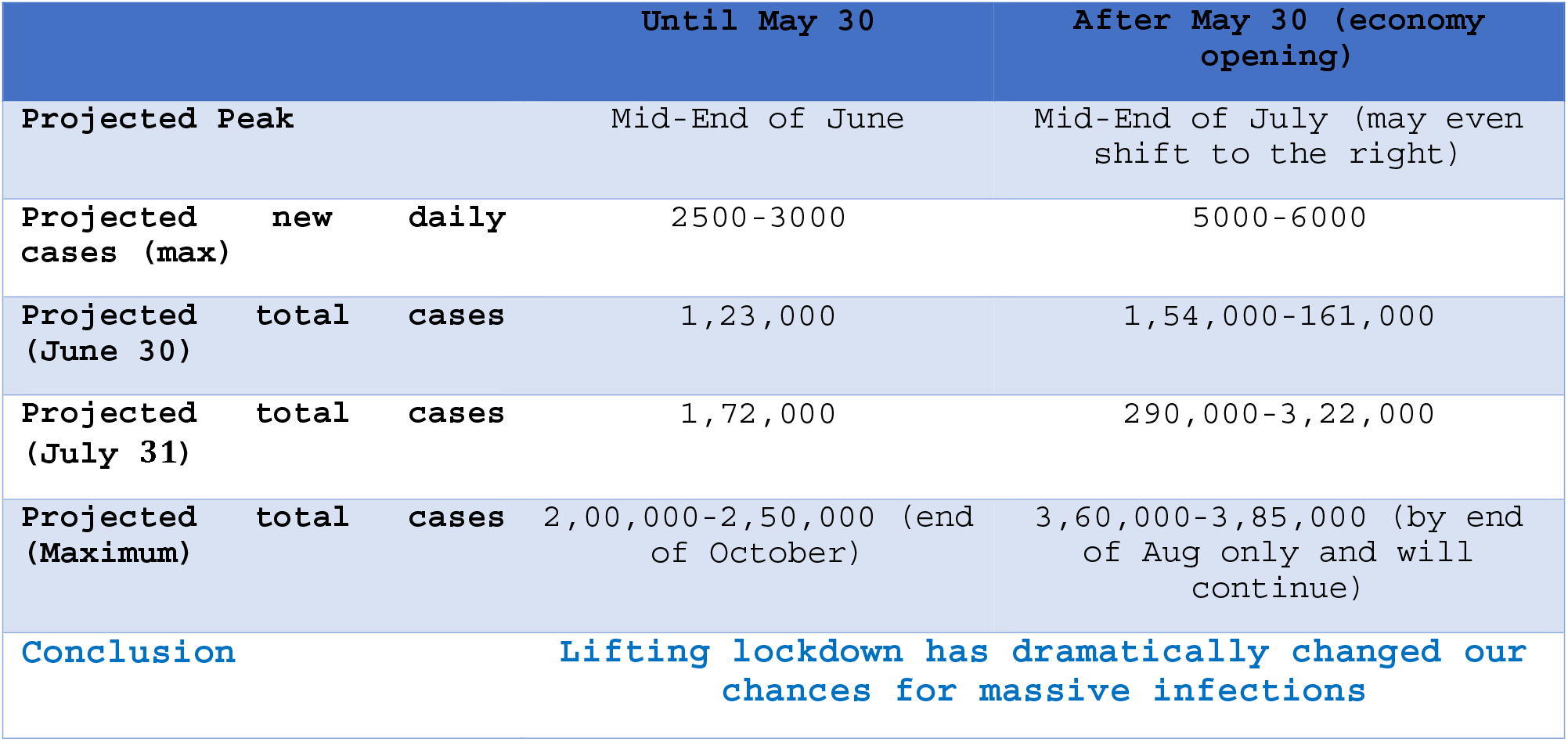

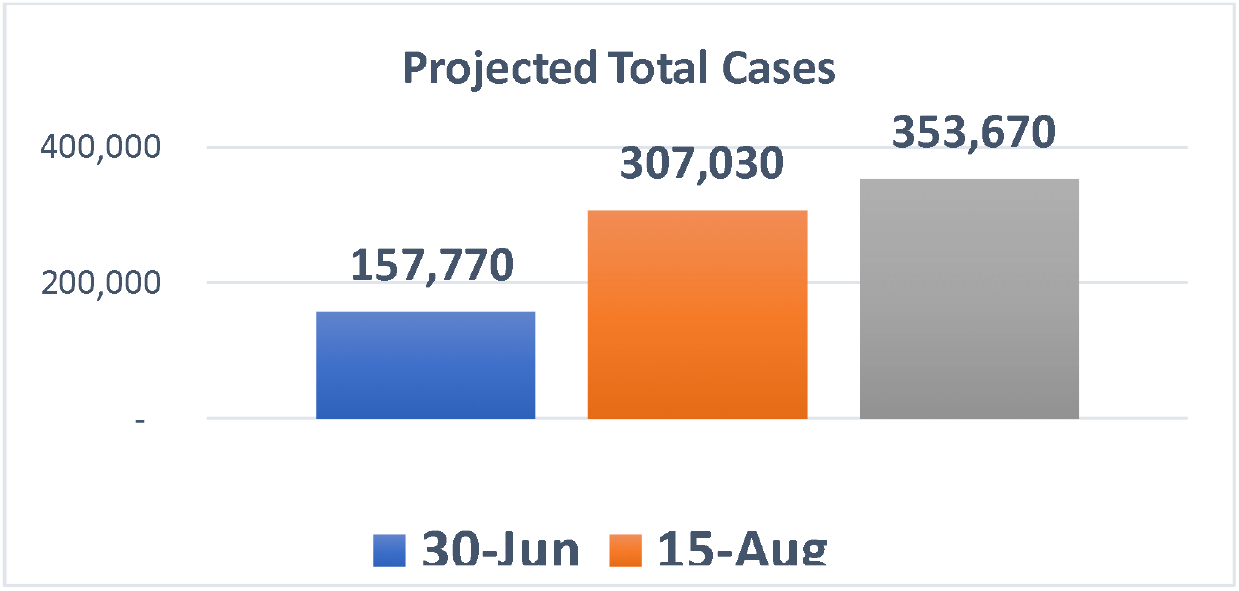

### F. Caution

New few days are very crucial. Infection has grown drastically after economy has opened. We have only 15 days after lockdown lifting, so complete picture yet to be seen.

### G. Conclusion

Bangladesh’s condition is now in very risky. If social distancing is maintained and people do not follow the precautionary measures, Bangladesh can next epic-center of the world like Brazil or Russia, if not the USA.

## Footnotes

1 The government declared a general holiday/ leave instead of calling it ‘lockdown’ or ‘shutdown’. However the objectives were the same—to tame the spread of the virus.

2 Technical Team: Shafiun Nahin Shimul, PhD, Institute of Health Economics, University of Dhaka & member Soho-Joddha Mofakhar Hussain, PhD, University of Toronto, Canada & member Soho-Joddha Dr. Abu Jamil Faisel, Public Health Expert Advisor, DGHS, MOHFW & member Soho-Joddha Syed Abdul Hamid, PhD, Institute of Health Economics, University of Dhaka & member Soho-Joddha

3 for up to date projection and more results visit https://sites.google.com/site/shafiunihe/recent-work-oncovid-19

